# Real-Time Glycaemic and Metabolic Adaptation During Unsupplemented Spiritual Fasting up to 30 Days: A Self-Controlled Observational Study

**DOI:** 10.64898/2026.07.20.26358472

**Authors:** Swapnil Prakash, Neha Shekhawat, Ojas Bardiya, Rahul Garg, Vaibhav Tripathi, Suruchi Fialoke

**Author notes:** Co-senior authors (Vaibhav Tripathi, Suruchi Fialoke). **Author Contributions** Supervision: S.F., V.T. Concept and design: S.F., V.T., R.G., S.P. Acquisition of data: N.S., S.F. Analysis and interpretation of data: N.S., O.B., S.F., V.T. Drafting of the manuscript: S.P., S.F., V.T. Medical supervision and safety monitoring: S.P.

## Abstract

**Background:** Prolonged unsupplemented spiritual fasting (USF), complete caloric abstinence motivated by spiritual practice, is undertaken by many communities worldwide, yet its physiological consequences remain poorly characterized. No prior study has documented continuous real-time monitoring during free-living fasting beyond 10 days.

**Methods:** We conducted a self-controlled observational study of 23 experienced Jain practitioners undergoing USF. Seventeen completed at least 8 days of fasting (11 completed eight days, six continued to 30 days); six discontinued early. Continuous glucose monitoring (CGM) and blood biomarkers at baseline (Day-0), post-fasting (Day-9), and 60-day follow-up (Day-69) were obtained. Mood was assessed daily using PANAS. Within-participant comparisons used both parametric and non-parametric t-tests.

**Results:** CGM revealed near-complete suppression of glycaemic variability within 24–48 hours of fasting onset, sustained throughout with no clinical hypoglycaemia in either cohort. Blood biomarkers showed transient perturbation during fasting — including rises in hepatic enzymes, bilirubin, uric acid, creatinine, and lipid fractions — broadly reversible by follow-up (Day-69). hsCRP rose during fasting then fell below baseline at follow-up (5.52±3.67 to 3.68±3.18 mg/L, p=0.034). HDL significantly rose above baseline (45.71±12.32 to 48.97±11.19, p=0.045) and LDL similarly declined below baseline (122.33±32.10 to 106.96±31.62 mg/dL, p=0.035); and then both significantly improved by follow-up. Thyroid axis suppression fully normalized by follow-up. Psychological wellbeing was maintained throughout.

**Conclusions:** Extended USF produces a safely reversible pattern of acute physiological adaptation with net cardiometabolic benefit. Absence of clinical hypoglycaemia during 30-day water-only USF, documented here for the first time with CGM, provides empirical grounding for future controlled trials.

## Introduction

Evolutionary biology suggests that human physiology is optimized for metabolic flexibility, an adaptation derived from intermittent nutrient availability, physiological demands, and environmental stressors. This homeostatic framework has evolved to pivot between fasted and postprandial states, exerting systemic effects that influence physiological, neurocognitive, and psychosocial domains. In recent times, a growing clinical consensus has emerged regarding the therapeutic potential of fasting in modulating cardiometabolic health, including obesity and diabetes^1–3^, oncological outcomes^4^, and autoimmune pathologies^5^. This interest has led to the formal categorization of various protocols, including intermittent fasting (IF), time-restricted eating (TRE), and medically supervised short-term fasting (STF)^6^. The physiological effects of intermittent and supplements-supported fasting are increasingly well characterized, yet the homeostatic consequences of prolonged fasting undertaken in free-living conditions, without a goal of weight reduction and instead motivated by spiritual goals, remain insufficiently defined.

This study examined a cohort of Jain householders practicing Unsupplemented Spiritual Fasting (USF) under the guidance of monks from the Jain-Terapanth tradition. Of 23 enrolled practitioners during Chaturmas (a sacred four-month holy period in Jainism, spanning the monsoon season) in Ahmedabad, 17 completed an 8-day fast, of which six continued for atleast 30 days. This population provides a unique model for clinical investigation owing to strict, uniform adherence to complete caloric abstinence without supplements, medications, or stimulants while maintaining normal physical and cognitive activities^7^. The biochemical signature of prolonged fasting is characterized by dynamic, biphasic shifts. Acute-phase responses typically reflect rapid mobilization of metabolic substrates and a transient increase in hormetic inflammatory signaling^8^. Conversely, longitudinal data suggest distinct homeostatic trajectories that result in sustained improvements in cardiovascular risk profiles^9^. Pilot investigations into spiritual fasting traditions corroborate these physiological shifts while consistently highlighting a distinct elevation in subjective well-being and positive affect following the fast^10^.

While conventional biochemical snapshots provide valuable insights into metabolic transitions, a significant gap exists regarding individualized, real-time metrics in prolonged fasting durations^7,11–13^. This study aims to address this deficit by integrating Continuous Glucose Monitoring (CGM)^14,15^ and Heart Rate Variability (HRV)^16^ to delineate the individualized autonomic and glycemic responses to USF. Psychological state was evaluated using the Positive and Negative Affect Schedule (PANAS) to quantify affective changes across the fasting period.

By corroborating these metrics with traditional metabolic markers including anthropometric, hepatic, renal, metabolic, inflammatory, and endocrinological profiles, this study provides a comprehensive perspective on the human capacity for homeostatic adaptation during USF.

## Methods

This study adopted a self-controlled observational design to investigate the physiological, psychological, and spiritual effects of USF in experienced practitioners. Each participant served as their own control, with repeated measurements collected across four distinct phases: a 4-day baseline period (Day −3 to Day 0), the unsupplemented spiritual fast (USF) phase (either 8 days or 30 days), a 4-day post-fasting recovery period, and a long-term follow-up conducted at day-69 (D69) after fasting began.

### Participants and Recruitment

We recruited participants through Jain fasting communities and spiritual organizations. Eligible participants were 18 to 70 years of age and had a body-mass index (BMI) above 18.5. Participants were excluded if they had uncontrolled metabolic, cardiovascular, autoimmune, hepatic, renal, or psychiatric conditions; a history of serious fasting-related complications (including Recurrent syncopes or life-threatening dehydration); or were pregnant or breastfeeding. Of 23 enrolled participants, 17 completed atleast 8-day USF, including 6 completing at least 30 days; 6 discontinued during the fasting phase. All subsequent analyses were done in the cohort of the 17 subjects (10 Female, 7 Male) completing at least 8 days USF. Male participants had a mean age of 50.4 years (range 34–63). Female participants (n=10) had a mean age of 45.6 years (range 33–54). Male participants had a mean BMI of 30.07 (range 25.86–34.09). Female participants had a mean BMI of 29.71 (range 23.82–37.50). All participants reported being vegetarians, as is common in Jainism.

### Ethics and Safety Monitoring

All participants provided written informed consent. The study was approved by the Institutional Ethics Committee of the Indian Institute of Technology, Gandhinagar. Participants were monitored under continuous medical supervision throughout the fasting phase. Predefined criteria for discontinuation included persistent dizziness, hypotension, critically low blood glucose, worsening of preexisting conditions, or voluntary withdrawal. Participants who discontinued were medically supervised during the recovery protocol; all data collected before withdrawal were retained for analysis.

### Assessments

Blood biomarker testing (Table 1) was performed at baseline (D0), immediately following the fasting phase (D9), and at long-term follow-up (D69). Continuous glucose monitoring (CGM; Sinocare) was conducted across all three periods^17^. Heart rate data were collected using the Polar H10 sensor or Polar Verity Sense. Daily Mood and affect were assessed using the Positive and Negative Affect Schedule (PANAS)^18^ along with sleep and meditation logs using structured pen-and-paper formats. During the fasting phase, daily assessments included blood pressure, pulse, oxygen saturation, body weight, and body-fat percentage; however, these measures were not obtained for all participants owing to practical constraints arising from participants being located at multiple sites. Following completion of the fasting phase, semi-structured interviews were conducted using predefined questions, audio-recorded, transcribed, anonymized, and stored on secure password-protected servers. Participant identifiers were removed prior to analysis.

**Table 1.** Values are expressed as mean ± SD. P-values are reported for two pairwise comparisons — baseline vs. post-fasting (D0–D9) and baseline vs. follow-up (D0–D69); derived from non-parametric, Wilcoxon signed-rank tests. Only participants with complete data at both timepoints of each comparison were included. Biomarkers are grouped by physiological system. hsCRP values reported as >10 mg/L were capped at 10 for analysis. Bold p-values indicate statistical significance at p<0.05.

| Biomarker Name | Baseline (D0) | Post (D9) | Followup (D69) | Baseline - Post |  | Baseline - Followup |  |
| --- | --- | --- | --- | --- | --- | --- | --- |
|  | Mean ± SD | Mean ± SD | Mean ± SD | Wilcoxon W | p-value | Wilcoxon W | p-value |
| Inflammatory and Cardiovascular Risk Marker |  |  |  |  |  |  |  |
| hsCRP | 5.52 ± 3.67 | 7.93 ± 2.63 | 3.68 ± 3.18 | 10 | 0.0076 | 27 | 0.034 |
| Liver Function Markers |  |  |  |  |  |  |  |
| ALT | 21.19 ± 9.26 | 26.96 ± 14.64 | 19.32 ± 8.89 | 40 | 0.0887 | 48 | 0.1901 |
| AST | 21.49 ± 5.04 | 32.59 ± 11.62 | 22.97 ± 8.36 | 12 | **0.0011** | 60 | 0.6791 |
| ALP | 73.45 ± 23.06 | 83.08 ± 27.48 | 81.47 ± 25.09 | 15 | **0.0021** | 21 | **0.0067** |
| S. Albumin | 4.18 ± 0.35 | 4.56 ± 0.21 | 3.72 ± 0.19 | 11 | **0.0054** | 1 | **<0.0001** |
| S. Globulin | 2.65 ± 0.53 | 3.30 ± 0.53 | 2.81 ± 0.41 | 2 | **<0.0001** | 48.5 | 0.1901 |
| A:G Ratio | 1.66 ± 0.44 | 1.42 ± 0.24 | 1.35 ± 0.21 | 20 | **0.013** | 10 | **0.0027** |
| GGTP | 19.46 ± 5.80 | 20.10 ± 5.44 | 18.22 ± 4.75 | 60.5 | 0.4586 | 55 | 0.3289 |
| S. Bilirubin<br>Direct | 0.11 ± 0.04 | 0.26 ± 0.07 | 0.12 ± 0.03 | 0 | **<0.0001** | 56.5 | 0.5513 |
| S. Bilirubin<br>Indirect | 0.42 ± 0.11 | 0.93 ± 0.30 | 0.44 ± 0.11 | 0 | **<0.0001** | 52 | 0.4077 |
| S. Bilirubin<br>Total | 0.53 ± 0.14 | 1.19 ± 0.37 | 0.55 ± 0.14 | 0 | **<0.0001** | 56.5 | 0.3529 |
| **Kidney Function Markers and Electrolytes** |  |  |  |  |  |  |  |
| S. Urea | 20.48 ± 5.64 | 35.82 ± 17.93 | 18.20 ± 7.66 | 3.5 | **0.0001** | 32 | **0.0348** |
| BUN | 9.57 ± 2.64 | 16.73 ± 8.37 | 8.50 ± 3.58 | 18.5 | **0.0103** | 60 | 1 |
| S. Creatinine | 0.73 ± 0.16 | 0.92 ± 0.20 | 0.63 ± 0.11 | 0 | **<0.0001** | 5 | **0.0002** |
| eGFR | 104.77 ±<br>12.05 | 89.12 ± 15.41 | 112.18 ± 8.05 | 0 | **<0.0001** | 6.5 | **0.0002** |
| S. Sodium | 138.21 ± 2.22 | 136.42 ± 2.98 | 139.63 ± 1.69 | 26 | **0.015** | 24 | **0.011** |
| S. Potassium | 3.96 ± 0.38 | 3.84 ± 0.49 | 3.96 ± 0.34 | 35 | 0.0879 | 68 | 1 |
| S. Chloride | 101.81 ± 2.11 | 96.22 ± 3.08 | 103.46 ± 1.32 | 0 | **<0.0001** | 13 | **0.0044** |
| S. Phosphorus | 4.37 ± 0.55 | 4.77 ± 1.08 | 4.54 ± 0.53 | 46 | 0.1594 | 52 | 0.2633 |
| S. Calcium | 9.05 ± 0.29 | 9.70 ± 0.30 | 8.95 ± 0.31 | 0 | **<0.0001** | 44.5 | 0.2242 |
| S. Uric Acid | 4.67 ± 1.26 | 12.19 ± 3.85 | 4.17 ± 1.19 | 1 | **<0.0001** | 29 | **0.0437** |
| **Lipid Profile** |  |  |  |  |  |  |  |
| Total<br>Cholesterol | 189.79 ±<br>39.60 | 256.68 ±<br>43.40 | 180.66 ± 37.45 | 0 | **<0.0001** | 55 | 0.3289 |
| HDL | 45.71 ± 12.32 | 43.28 ± 9.90 | 48.97 ± 11.19 | 41 | 0.0984 | 34.5 | **0.0448** |
| LDL | 122.33 ±<br>32.10 | 188.52 ±<br>38.13 | 106.96 ± 31.62 | 0 | **<0.0001** | 32 | **0.0348** |
| VLDL | 21.75 ± 9.03 | 24.88 ± 8.66 | 24.74 ± 14.42 | 27 | **0.0174** | 53 | 0.2842 |
| Triglyceride | 108.77 ±<br>45.15 | 124.39 ±<br>43.31 | 123.69 ± 72.09 | 27 | **0.0174** | 53 | 0.2842 |
| **Metabolic and Endocrine Functions** |  |  |  |  |  |  |  |
| Total T3 | 1.17 ± 0.24 | 0.65 ± 0.15 | 1.21 ± 0.36 | 0 | **0** | 75 | 0.9632 |
| Total T4 | 6.96 ± 1.39 | 8.52 ± 1.60 | 6.83 ± 1.57 | 0 | **0** | 63 | 0.5477 |
| TSH | 4.22 ± 2.16 | 2.71 ± 1.84 | 4.21 ± 2.72 | 8 | **0.0004** | 69 | 0.7467 |
| Glucose<br>Fasting | 91.81 ± 18.34 | 71.75 ± 9.26 | 85.66 ± 11.42 | 20 | **0.0056** | 51 | 0.2435 |

### Analysis

#### Glucose data Preprocessing

Raw CGM files were merged and records with missing timestamps and non-numeric values were removed. The dataset was restricted to baseline (D−3 to D0), fasting, and post-fasting phases, with all dates expressed as study days relative to fasting onset (D0). For the stacked individual traces (Figure 2A, 2B), glucose values were z-scored and plotted with fixed vertical offsets. For hourly group-level analysis (Figure 2C), raw glucose values were binned into 24 hourly windows per study day and averaged across participants, with daytime (06:00–18:00) and nighttime (18:00–06:00) distinguished by colour. Group-level mean ± SD was computed for each bin. Subject-level plots of raw CGM data are available in Supplementary Information.

**Figure 1.**
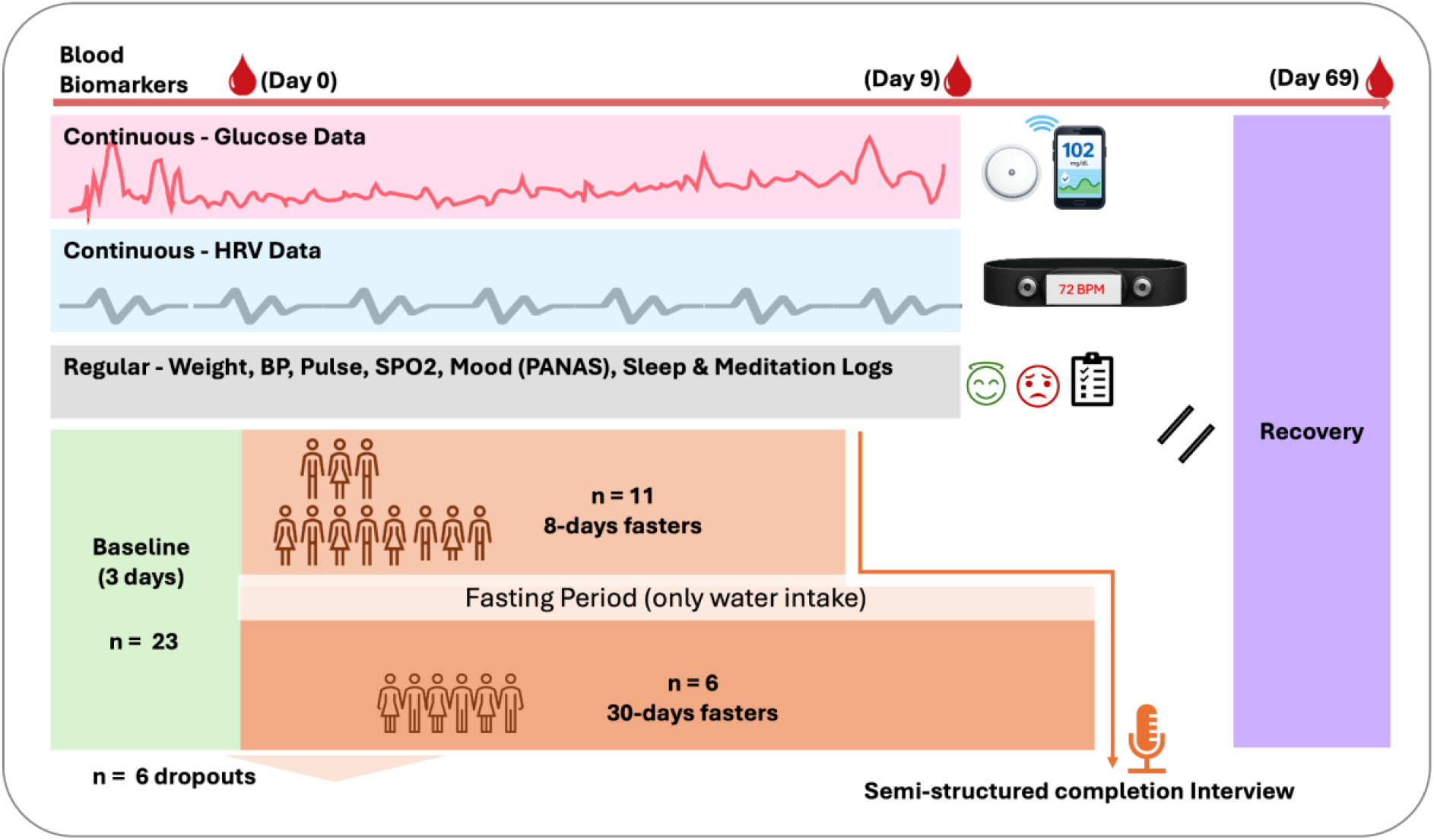
Study schematic. Participants (n=23) completed a 3-day baseline period followed by an unsupplemented water-only fasting phase, during which 11 participants fasted for eight days, and six continued to 30 days; six individuals discontinued early. Continuous glucose and heart-rate variability monitoring was performed throughout, along with regular assessments of weight, blood pressure, pulse, oxygen saturation, mood (PANAS), sleep, and meditation logs. Blood biomarkers were obtained on days 0, 9, and 69. A semi-structured interview was conducted at completion. All participants then entered a supervised recovery period. Analyses were restricted to the 17 participants completing at least 8 days of USF.

**Figure 2.**
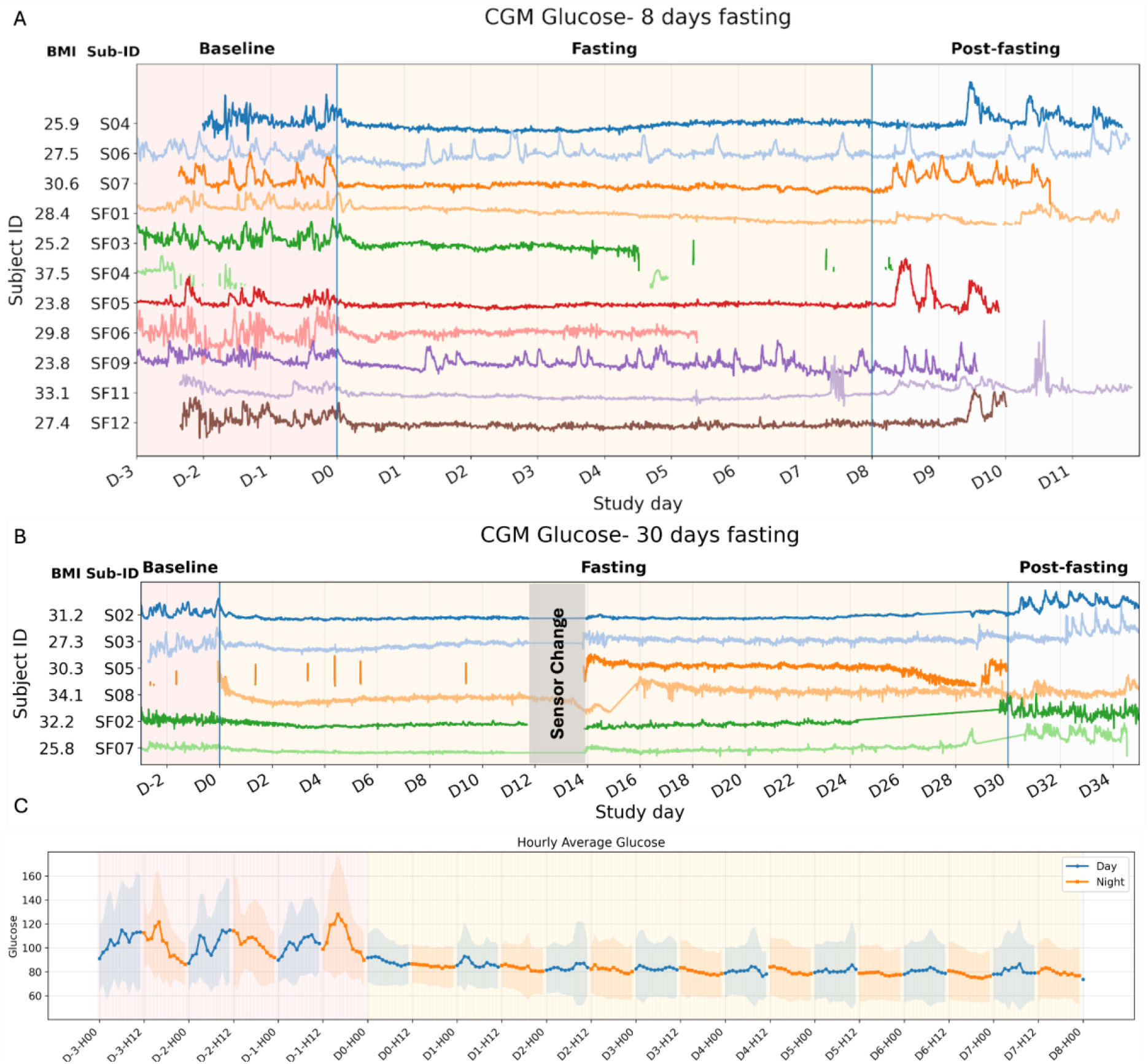
Continuous glucose monitoring (CGM) traces across the study period for all 17 participants including(A)individual CGM traces for 8-day fasters (n=11) and (B) 30-day fasters (n=6). Each colored trace represents one participant; BMI and subject ID are shown on the left (subject IDs starting from “SF” represent female subjects). Vertical blue lines demarcate phase transitions. Background shading denotes study phases: pink=baseline, yellow=fasting, white=post-fasting. Gaps in traces reflect sensor dropout and a planned sensor change at approximately Day 14 is annotated in (B). (C) Group-level mean hourly glucose averaged across the 8-day fasting cohort, with shaded bands representing ±1 SD. Blue indicates daytime (06:00–18:00) and orange indicates nighttime (18:00–06:00).

#### Blood biomarker analysis

For each biomarker, paired t-tests (parametric, assumes normality) and Wilcoxon signed-rank tests (non-parametric, rank-based) were conducted for two comparisons: baseline vs. post-fasting (D0–D9) and baseline vs. follow-up (D0–D69), including only participants with complete pairs at each timepoint. Results were concordant across both methods for all primary findings. Biomarker distributions were visualized as split violin plots with overlaid individual data points, with male (n=7) and female (n=10) participants shown on left and right respectively.

#### Heart rate variability (HRV)

was recorded continuously using the Polar H10 chest strap in four of the six thirty-day fasters and two eight-day fasters. HRV metrics were computed using NeuroKit2 on recordings exceeding two minutes; parameters extracted included RMSSD, SDNN, LF/HF ratio, and log(HF)^19^. Average heart rate was computed per sample. Usable HRV data were obtained from four thirty-day fasters, yielding 554 samples (mean 138.5 per participant); data from 8-day fasters were excluded owing to high signal noise.

#### Other Variables

Daily variables (weight, blood pressure, pulse rate, SpO_2_, and PANAS scores) were analyzed using data from a defined window of baseline (D-3 to D0) and post-fasting (D5 to D8), due to inconsistent daily recordings. When multiple values were available, the most recent measurement in each window was selected. Only participants with data at both time points were included. For each variable, mean ± SD was calculated at baseline and post-fasting, and changes were assessed using Wilcoxon signed-rank test statistics, with significance determined by p-values.

## Results

### Continuous Glucose Findings

CGM revealed a consistent and visually striking suppression of glycaemic variability upon entry into the fasting phase across all 17 participants (Figure 2). During the baseline period, glucose traces exhibited characteristic postprandial oscillations, with frequent excursions reflecting normal meal-related fluctuations. Following the onset of fasting (left blue vertical line), glucose levels declined rapidly and stabilised at markedly lower values, with near-complete attenuation of the oscillatory pattern observed in virtually all subjects, reflecting the depletion of glycogen stores and a metabolic shift toward gluconeogenesis and ketone utilisation as primary energy substrates. This suppression was sustained throughout the fasting phase, with glucose traces remaining flat and low across both the 8-day and 30-day fasting groups for most subjects. All 17 subjects confirmed no intake of any food, electrolyte or medicine apart from plain water throughout their respective fasting window. Among the 30-day fasters (Figure 2B), this flattened, low-glucose profile persisted for the extended fasting duration with no evidence of clinical hypoglycaemia, suggesting stable counter-regulatory adaptation over remarkably prolonged USF. Upon refeeding (right blue vertical line), glucose variability promptly resumed in the majority of participants, with traces returning to an oscillatory pattern consistent with reintroduction of caloric intake. Notably, some subjects (e.g., S04, SF05; Figure 2A) exhibited more pronounced post-refeeding excursions relative to their own baseline, while others showed a comparatively muted refeeding response, pointing to inter-individual heterogeneity in glycaemic recovery. SF03, SF04 (2A) and S05 (2B), have notable data gaps during the mid-fasting period, reflecting sensor dropout rather than physiological discontinuity. Among the 30-day fasters (Figure 2B), the flattened low-glucose profile persisted across the full extended duration with no evidence of progressive hypoglycaemic deterioration, and group-level hourly analysis (Figure 2C) confirmed convergence of daytime and nighttime glucose means to similarly suppressed levels throughout the fasting phase, with divergence resuming upon refeeding.

### Blood Biomarker Findings

For blood biomarkers, changes were assessed using Wilcoxon signed-rank tests and corresponding p-values are reported in Table 1; paired t-tests were performed in parallel (details in Supplementary). Results were concordant across both methods for all primary findings. Biomarker profiles across the three timepoints revealed a consistent pattern of acute physiological perturbation during fasting that was broadly reversible by 60-day follow-up, with several markers showing net improvement relative to baseline.

hsCRP rose transiently during fasting (5.52±3.67 to 7.93±2.63 mg/L, p=0.008) and fell below baseline at follow-up (3.68±3.18 mg/L, p=0.03), indicating a net anti-inflammatory effect (Figure 3A). Values reported as >10 mg/L were capped at 10 for analysis, likely underestimating the fasting-phase rise. AST rose significantly during fasting (p=0.001) and normalized by follow-up; ALT showed a non-significant trend (p=0.089). Bilirubin fractions increased sharply (all p<0.0001), consistent with transiently reduced hepatic clearance, and fully resolved by followup (p>0.1). Serum albumin rose during fasting (4.18±0.35 to 4.56±0.21 g/dL, p=0.005), likely reflecting haemoconcentration, then fell below baseline at follow-up (3.72±0.19 g/dL, p<0.0001) (Figure 3B).

**Figure 3.**
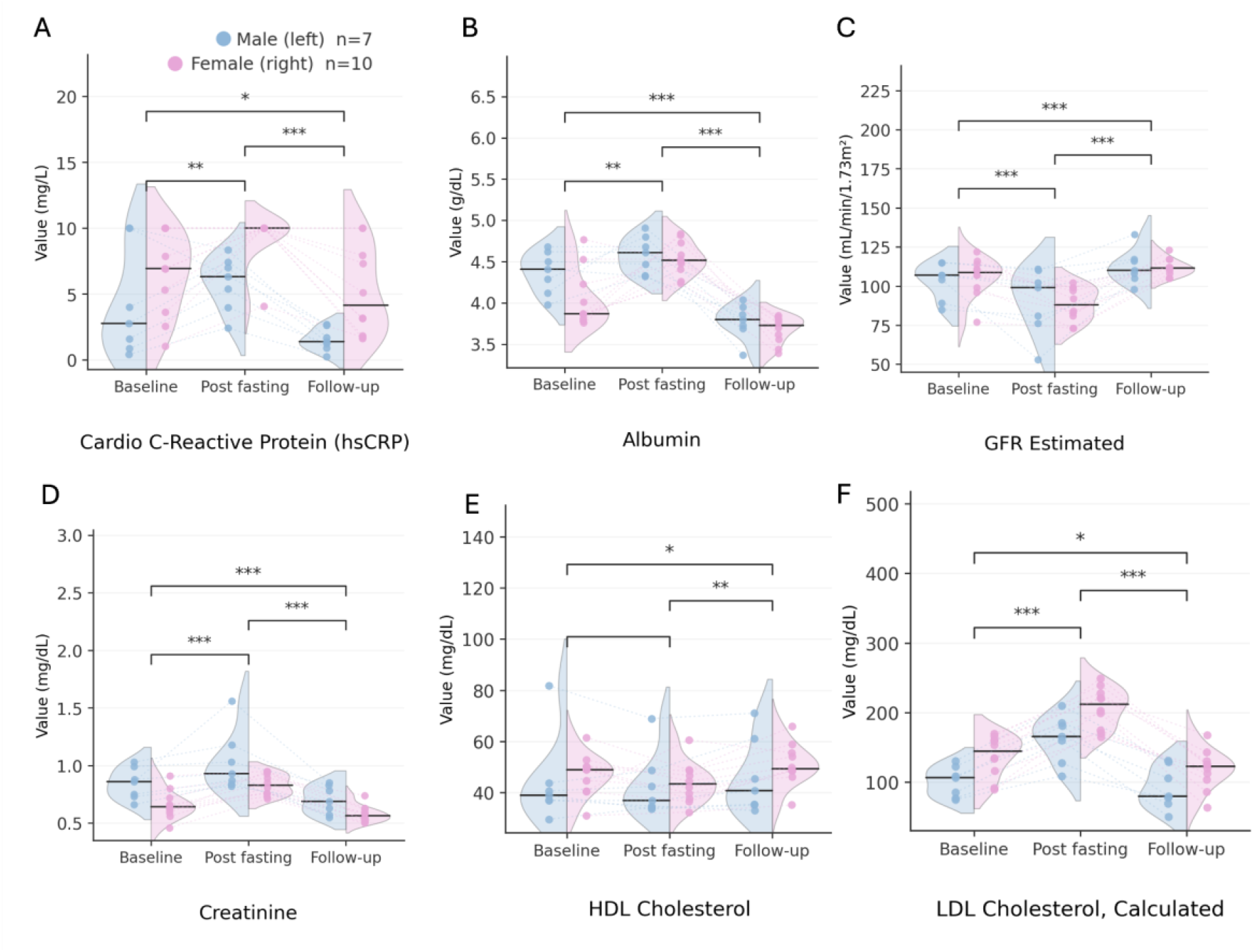
Selected Blood Biomarkers Across Baseline, Post-Fasting, and Follow-Up Timepoints. Split violin plots show the distribution of values for male (blue, left; n=7) and female (pink, right; n=10) participants at each timepoint. Horizontal lines within each violin denote the median. Individual data points are overlaid. Brackets with p-values indicate pairwise comparisons between timepoints obtained via paired t-tests. (A) hsCRP rose significantly during fasting (substantially more for females) and fell below baseline at follow-up, suggesting a net anti-inflammatory effect. (B) Serum albumin rose during fasting, consistent with haemoconcentration, and declined below baseline at follow-up; this post-recovery suppression was more pronounced in female participants. (C) eGFR declined during fasting and recovered above baseline at follow-up, consistent with transient haemoconcentration rather than intrinsic renal injury. (D) Serum creatinine rose during fasting and fell below baseline at follow-up, mirroring the eGFR trajectory. (E) HDL cholesterol rose above baseline at follow-up, suggesting a net favourable lipid effect. (F) LDL cholesterol rose markedly during fasting, reflecting hepatic lipid mobilisation, and fell below baseline at follow-up, indicating durable metabolic benefit.

Serum uric acid surged during fasting (4.67±1.26 to 12.19±3.85 mg/dL, p<0.0001), consistent with nucleotide turnover and reduced renal clearance in a ketotic state, and improved by follow-up (4.17±1.19, p=0.044)^20^. Urea and BUN approximately doubled (both p<0.01), consistent with gluconeogenic protein catabolism and finally improved at followup. Creatinine rose and eGFR declined during fasting (both p<0.001), likely reflecting haemoconcentration rather than intrinsic renal injury; both improved significantly below baseline by follow-up (both p<0.001) (Figure 3C and 3D). Electrolytes remained within physiologically stable ranges throughout.

Total cholesterol and LDL rose markedly during fasting (both p<0.0001), reflecting hepatic lipid mobilisation. At follow-up, LDL fell below baseline (106.96±31.62 vs. 122.33±32.10 mg/dL, p=0.035) and HDL rose above baseline (48.97±11.19 vs. 45.71±12.32, p=0.045). Fasting glucose declined during the fasting phase (91.81±18.34 to 71.75±9.26 mg/dL, p=0.005) and remained modestly below baseline at follow-up (85.66±11.42, p=0.20). The thyroid axis showed a characteristic adaptive response: T3 fell (p<0.0001), T4 rose (p<0.0001), and TSH declined (p<0.001), consistent with suppression of the hypothalamic-pituitary-thyroid axis to conserve energy^21^. All three parameters fully normalized by Day 69 (all p>0.50).

### Other Findings

Weight, BP, Pulse, SPO2 and PANAS and Heart rate data could not be obtained for the full cohort owing to participants being distributed across multiple sites and practical constraints during the monsoon period; self-reported measurements were not accepted. Among the 8 participants for whom in-person measurements were feasible, mean body weight decreased by 7.6 kg (81.19 ± 12.33 to 73.50 ± 8.64 kg, p = 0.007) and BMI fell by 2.8 units (29.85 ± 4.75 to 27.07 ± 3.85 kg/m^2^, p = 0.007) over 8 days of USF. Systolic blood pressure showed a clinically meaningful reduction of 10.4 mmHg (130.29 ± 18.22 to 119.86 ± 9.70 mmHg) that did not reach statistical significance in this limited sample (p = 0.2, n = 7); diastolic blood pressure, pulse, and oxygen saturation remained stable throughout (all p > 0.10), indicating preserved haemodynamic and respiratory integrity.

Aggregate positive affect (PANAS-P) was unchanged from baseline to post-fasting (39.25±13.25 vs. 38.63±8.88, p=0.74, n=8). Negative affect (PANAS-N) showed a trend toward reduction (14.88±4.58 vs. 12.38±1.19, p=0.17, n=8) that did not reach statistical significance, likely reflecting a floor effect given the low baseline negativity in this cohort. At the item level (Figure 4), negative affect items including Distressed, Upset, Hostile, Irritable, Jittery, and Afraid, were uniformly low at baseline and showed further reduction post-fasting, leaving little room for detectable change. Among positive affect items, post-fasting scores were approximately the same. Taken together, these findings indicate that psychological wellbeing was well-maintained across the fasting period, with no evidence of affective deterioration.

**Figure 4.**
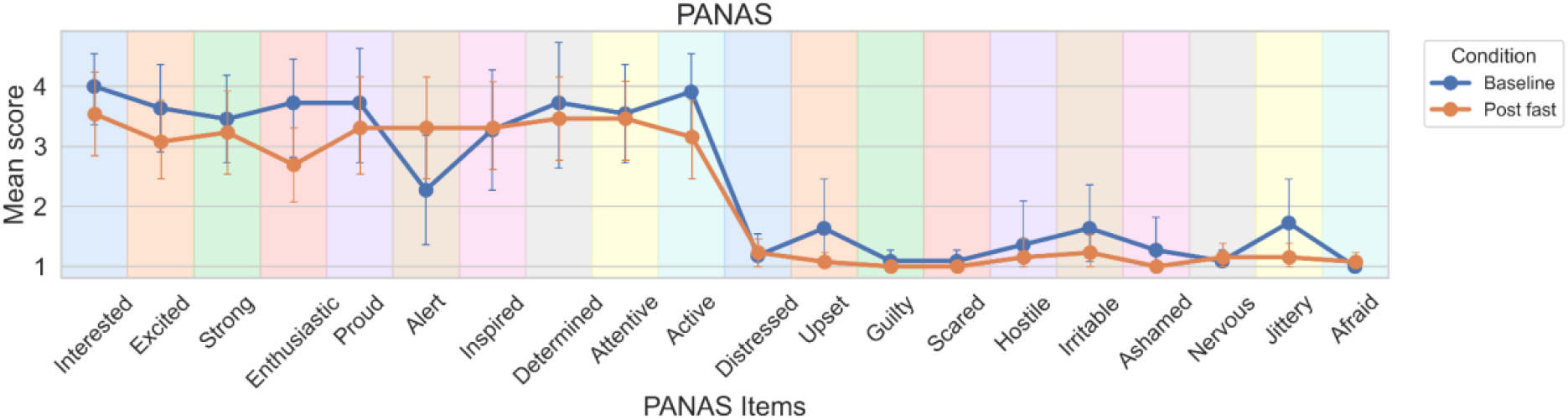
PANAS Item-Level Scores at Baseline and Post-Fasting. Mean scores (±95% CI) for all 20 PANAS items at baseline (blue) and post-fasting (orange). Items are arranged along the x-axis in order of positive affect (left, items 1–10: Interested through Active) followed by negative affect (right, items 11–20: Distressed through Afraid). Background shading distinguishes individual items for readability. Negative affect items were uniformly low at both timepoints, consistent with a floor effect. Positive affect items were broadly stable across the fasting period, with overlapping confidence intervals at all items.

Usable HRV data were obtained from 4 thirty-day fasters; data from 8-day fasters were excluded owing to signal noise. Pre-fasting recordings were limited to three samples from a single participant, precluding formal statistical comparison. Trends toward decreased heart rate, increased RMSSD and SDNN, and reduced LF/HF ratio during and after fasting were consistent with augmented parasympathetic drive, though none reached statistical significance given the limited baseline (Figure S3). Intra-day HRV patterns for representative participants are shown in Figure S4.

## Conclusion

This study provides the first continuous real-time characterization of glycaemic adaptation during extended unsupplemented spiritual fasting (USF) in free-living practitioners (n=6, 30 days fasters and n=11, 8-days fasters). Three principal findings emerge: sustained suppression of glycaemic variability throughout fasting with no clinical hypoglycaemia even at 30 days; a consistent pattern of acute physiological perturbation across hepatic, renal, lipid, and endocrine systems that was broadly reversible by 2-month follow-up; and preservation of psychological wellbeing throughout.

The CGM data are particularly instructive. Near-complete attenuation of postprandial glucose oscillations within the first 24–48 hours, followed by stable low-glucose profiles sustained for up to 30 days, corroborates the classical adaptive fasting response; glycogenolysis giving way to gluconeogenesis and ketogenesis as primary energy substrates^13^. Critically, the absence of progressive hypoglycaemic deterioration among both 8-day (n=11) and 30-day fasters (n=6), all maintaining normal physical and cognitive activity, suggests robust counter-regulatory competence not previously documented with continuous monitoring over this duration^11,15^. The transient elevation of hepatic enzymes, bilirubin, uric acid, creatinine, and lipid fractions is consistent with haemoconcentration and hepatic lipid mobilisation rather than organ injury, supported by complete normalization by 2 months. Bilirubin rise during fasting is a well-characterized phenomenon driven by competitive inhibition of hepatic clearance by circulating free fatty acids^8^. The initial fasting period revealed a hormetic inflammatory response: hsCRP rose transiently but fell significantly below baseline at follow-up, a pattern consistent with biphasic, baseline-dependent immune modulation described in larger fasting cohorts^22^, and with the sustained reductions in hsCRP and LDL cholesterol previously reported at six-week follow-up in medically supervised water-only fasting^9^. Several limitations apply: the self-controlled design precludes causal inference; and the exclusively Jain, experienced-practitioner (n=17) cohort limits generalizability. Overall, USF produced a mechanistically coherent, safely reversible pattern of acute adaptation with net cardiometabolic benefit at 60 days, providing empirical grounding for future prospective trials.

## Supporting information

Supplementary Results

## Data Availability

All data produced in the present study are available upon reasonable request to the authors.

## Acknowledgements

The authors gratefully acknowledge the spiritual guidance and blessings of Rev. Acharya Shri Mahashraman whose support made this study possible. We thank the volunteers and field monitors who assisted with data collection, and most importantly, the participants whose dedication and commitment to this demanding protocol made this research possible. Financial support was provided by Bhairon Lal Ratan Devi Bardiya Charitable Society.

## Notes

### Competing Interest Statement

The authors have declared no competing interest.

### Author Declarations

Institutional Ethics Committee (IEC) of Indian Institute of Technology, Gandhinagar gave ethical approval for this work.

