## Supplementary Results for "Real-Time Glycaemic and Metabolic Adaptation During Unsupplemented Spiritual Fasting up to 30 Days: A Self-Controlled Observational Study"

**Table S1.** Values are expressed as mean ± SD. P-values are reported for two pairwise comparisons — baseline vs. post-fasting (D0–D9) and baseline vs. follow-up (D0–D69); derived from paired t-tests (p-value, parametric). Only participants with complete data at both timepoints of each comparison were included. Biomarkers are grouped by physiological system. hsCRP values reported as >10 mg/L were capped at 10 for analysis. Bold p-values indicate statistical significance at p<0.05.

| Biomarker Name | Baseline (D0) | Post (D9) | Followup (D69) | Baseline - Post | | Baseline - Followup | |
|---|---|---|---|---|---|---|---|
| | Mean ± SD | Mean ± SD | Mean ± SD | T stat | p-value | T stat | p-value |
| **Inflammatory and Cardiovascular Risk Marker** | | | | | | | |
| hsCRP | 5.52 ± 3.67 | 7.93 ± 2.63 | 3.68 ± 3.18 | -3.55 | **0.0027** | 2.31 | **0.0347** |
| **Liver Function Markers** | | | | | | | |
| ALT | 21.19 ± 9.26 | 26.96 ± 14.64 | 19.32 ± 8.89 | -2.07 | 0.0554 | 0.69 | 0.4982 |
| AST | 21.49 ± 5.04 | 32.59 ± 11.62 | 22.97 ± 8.36 | -3.88 | **0.0013** | -0.65 | 0.5239 |
| ALP | 73.45 ± 23.06 | 83.08 ± 27.48 | 81.47 ± 25.09 | -4.08 | **0.0009** | -2.85 | **0.0115** |
| S. Albumin | 4.18 ± 0.35 | 4.56 ± 0.21 | 3.72 ± 0.19 | -3.86 | **0.0014** | 5.71 | **<0.0001** |
| S. Globulin | 2.65 ± 0.53 | 3.30 ± 0.53 | 2.81 ± 0.41 | -5.43 | **<0.0001** | -1.47 | 0.1596 |
| A:G Ratio | 1.66 ± 0.44 | 1.42 ± 0.24 | 1.35 ± 0.21 | 2.31 | **0.0347** | 2.87 | **0.0112** |
| GGTP | 19.46 ± 5.80 | 20.10 ± 5.44 | 18.22 ± 4.75 | -1.29 | 0.2162 | 1.07 | 0.2985 |
| S. Bilirubin Direct | 0.11 ± 0.04 | 0.26 ± 0.07 | 0.12 ± 0.03 | -9.00 | **<0.0001** | -0.75 | 0.4647 |
| S. Bilirubin Indirect | 0.42 ± 0.11 | 0.93 ± 0.30 | 0.44 ± 0.11 | -8.19 | **<0.0001** | -0.82 | 0.4248 |
| S. Bilirubin Total | 0.53 ± 0.14 | 1.19 ± 0.37 | 0.55 ± 0.14 | -8.59 | **<0.0001** | -0.89 | 0.3857 |
| **Kidney Function Markers and Electrolytes** | | | | | | | |
| S. Urea | 20.48 ± 5.64 | 35.82 ± 17.93 | 18.20 ± 7.66 | -3.74 | **0.0018** | 1.28 | 0.2202 |
| BUN | 9.57 ± 2.64 | 16.73 ± 8.37 | 8.50 ± 3.58 | -2.96 | **0.0018** | -0.18 | 0.2194 |
| S. Creatinine | 0.73 ± 0.16 | 0.92 ± 0.20 | 0.63 ± 0.11 | -4.68 | **0.0003** | 4.28 | **0.0006** |
| eGFR | 104.77 ± 12.05 | 89.12 ± 15.41 | 112.18 ± 8.05 | 4.76 | **0.0002** | -3.47 | **0.0032** |
| S. Sodium | 138.21 ± 2.22 | 136.42 ± 2.98 | 139.63 ± 1.69 | 2.65 | **0.0176** | -3.00 | **0.0084** |
| S. Potassium | 3.96 ± 0.38 | 3.84 ± 0.49 | 3.96 ± 0.34 | 1.34 | 0.1993 | -0.09 | 0.9326 |
| S. Chloride | 101.81 ± 2.11 | 96.22 ± 3.08 | 103.46 ± 1.32 | 6.63 | **<0.0001** | -3.42 | **0.0035** |
| S. Phosphorus | 4.37 ± 0.55 | 4.77 ± 1.08 | 4.54 ± 0.53 | -1.59 | 0.1324 | -1.37 | 0.1905 |
| S. Calcium | 9.05 ± 0.29 | 9.70 ± 0.30 | 8.95 ± 0.31 | -7.56 | **<0.0001** | 1.40 | 0.1795 |
| S. Uric Acid | 4.67 ± 1.26 | 12.19 ± 3.85 | 4.17 ± 1.19 | -11.17 | **<0.0001** | 2.24 | **0.0395** |
| **Lipid Profile** | | | | | | | |
| Total Cholesterol | 189.79 ± 39.60 | 256.68 ± 43.40 | 180.66 ± 37.45 | -9.82 | **<0.0001** | 1.18 | 0.2545 |
| HDL | 45.71 ± 12.32 | 43.28 ± 9.90 | 48.97 ± 11.19 | 1.25 | 0.2305 | -1.93 | 0.0710 |
| LDL | 122.33 ± 32.10 | 188.52 ± 38.13 | 106.96 ± 31.62 | -11.88 | **<0.0001** | 2.46 | 0.0254 |
| VLDL | 21.75 ± 9.03 | 24.88 ± 8.66 | 24.74 ± 14.42 | -1.93 | 0.0716 | -1.07 | 0.2997 |
| Triglyceride | 108.77 ± 45.15 | 124.39 ± 43.31 | 123.69 ± 72.09 | -1.93 | 0.0716 | -1.07 | 0.2997 |
| **Metabolic and Endocrine Functions** | | | | | | | |

| | | | | | | | |
|---|---|---|---|---|---|---|---|
| Total T3 | 1.17 ± 0.24 | 0.65 ± 0.15 | 1.21 ± 0.36 | 11.12 | **<0.0001** | -0.36 | 0.7243 |
| Total T4 | 6.96 ± 1.39 | 8.52 ± 1.60 | 6.83 ± 1.57 | -5.54 | **<0.0001** | 0.67 | 0.5137 |
| TSH | 4.22 ± 2.16 | 2.71 ± 1.84 | 4.21 ± 2.72 | 4.36 | **0.0005** | 0.01 | 0.9883 |
| Glucose Fasting | 91.81 ± 18.34 | 71.75 ± 9.26 | 85.66 ± 11.42 | 3.79 | **0.0018** | 1.34 | 0.2008 |

**Table S2.** Results of normality testing and statistical test for each outcome variable, including physiological measures ($SpO_2$, pulse rate, systolic and diastolic blood pressure), psychological measures (PANAS Positive and Negative Affect scores), and anthropometric measures (weight and BMI). The table presents sample size (*N*), Shapiro–Wilk test results, paired *t*-test statistics and *p*-values, Wilcoxon signed-rank test statistics and *p*-values. Shapiro–Wilk test *p*-values were used to assess the normality of change scores for each outcome variable.

| Variable | N | Shapiro_p | Paired_t_stat | Paired_t_p value | Wilcoxon_stat | Wilcoxon_p |
|---|---|---|---|---|---|---|
| SPO2 | 6 | 0.0221 | -1.3868 | 0.2242 | 4 | 0.2188 |
| Pulse | 7 | 0.1118 | -0.3528 | 0.7363 | 10.5 | 0.5781 |
| Systolic BP | 7 | 0.5251 | 1.7452 | 0.1316 | 6 | 0.2188 |
| Diastolic BP | 7 | 0.8634 | 0.2797 | 0.7891 | 13 | 0.9375 |
| PANAS Positive | 8 | 0.7777 | 0.1403 | 0.8924 | 14.5 | 0.7422 |
| PANAS Negative | 8 | 0.3026 | 1.6938 | 0.1341 | 6 | 0.1732 |
| Weight | 8 | 0.362 | 4.0382 | 0.0049 | 0 | 0.0078 |
| BMI | 8 | 0.3395 | 4.1547 | 0.0043 | 0 | 0.0078 |

**Figure S1. Individual continuous glucose monitoring (CGM) profiles during an 8-day fasting.** Raw glucose concentrations (mg/dL) are plotted across study days. Shaded regions represent the baseline (pink), fasting (yellow), and post-fasting/refeeding (blue-grey) phases.

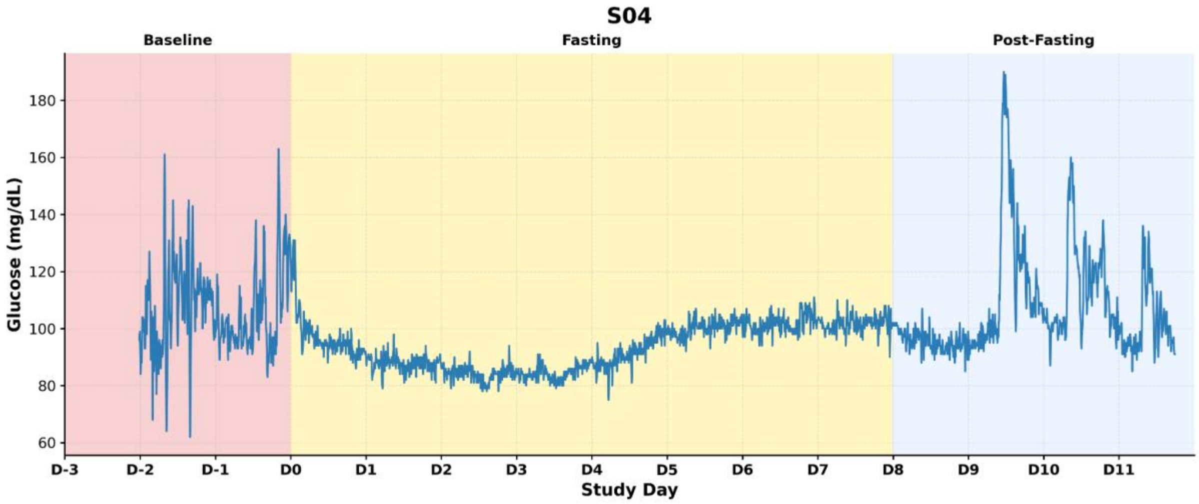


*Figure S1.1 Glucose time-series profile of Subject 04*

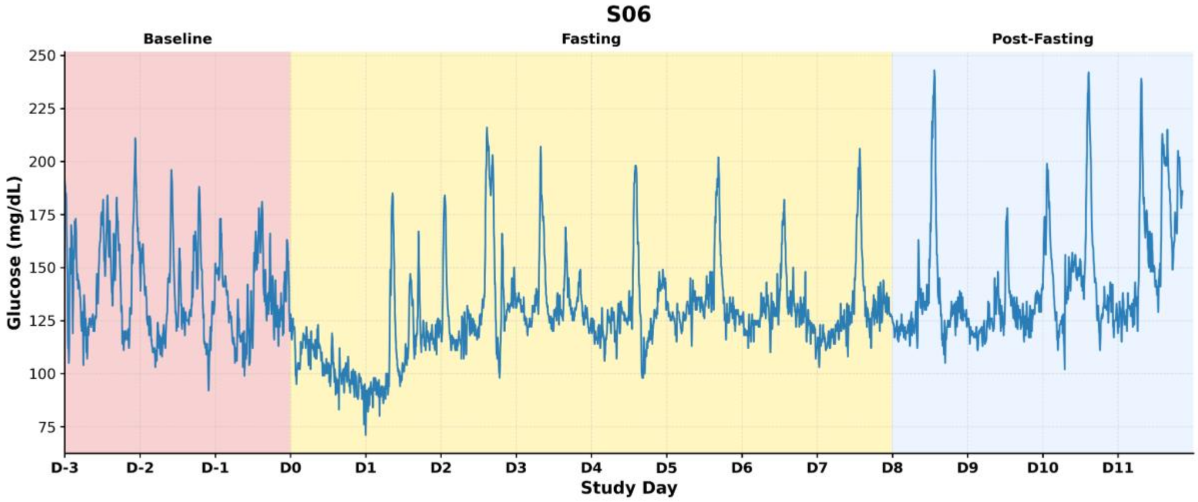


*Figure S1.2 Glucose time-series profile of Subject 06*

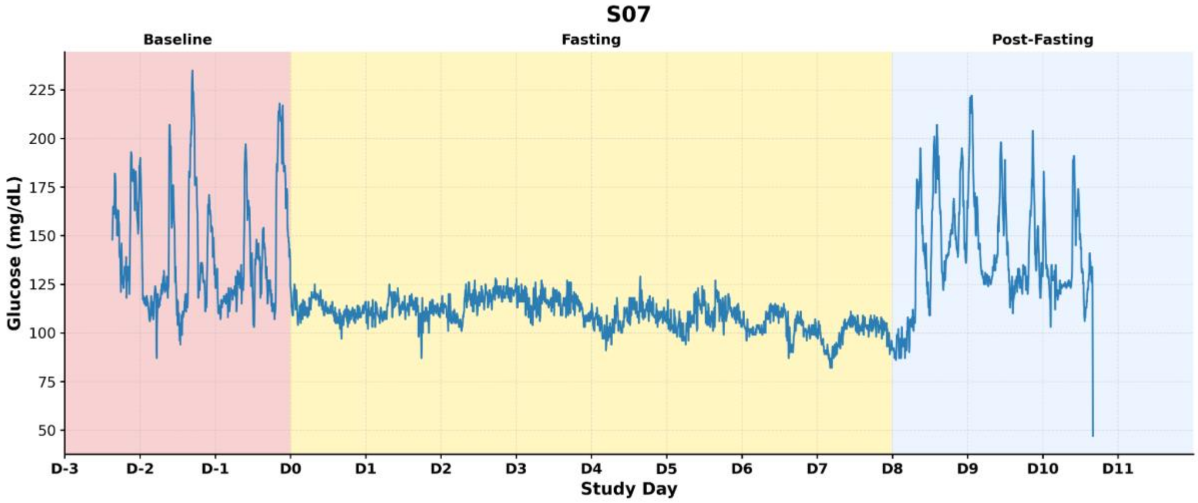


*Figure S1.3 Glucose time-series profile of Subject 07*

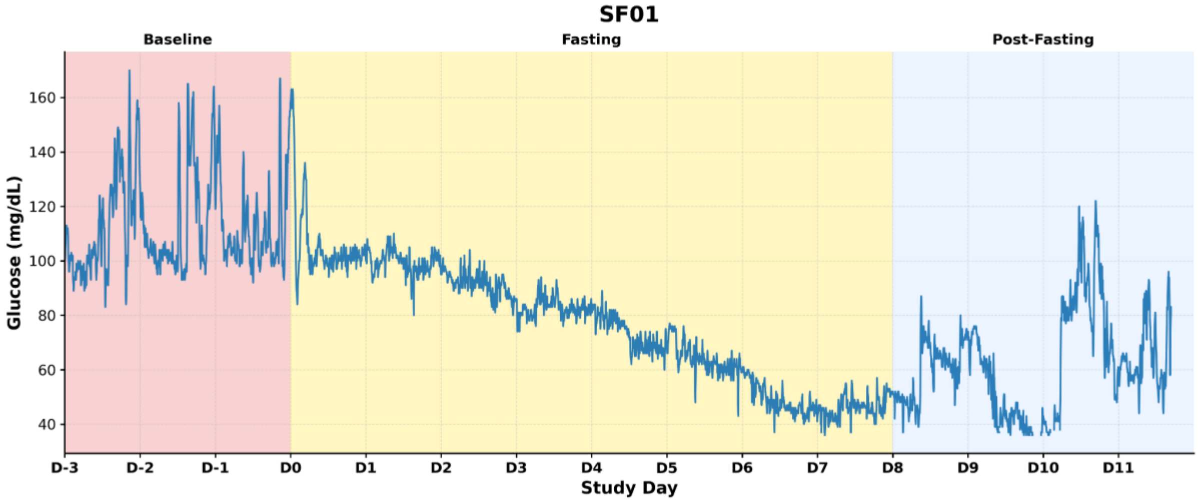


*Figure S1.4 Glucose time-series profile of Subject F01*

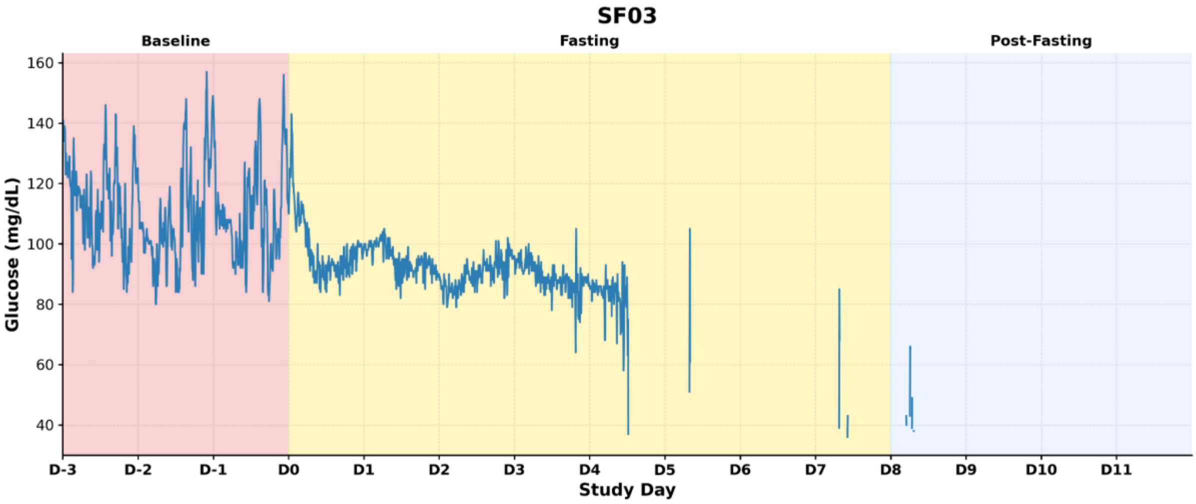


*Figure S1.5 Glucose time-series profile of Subject F03*

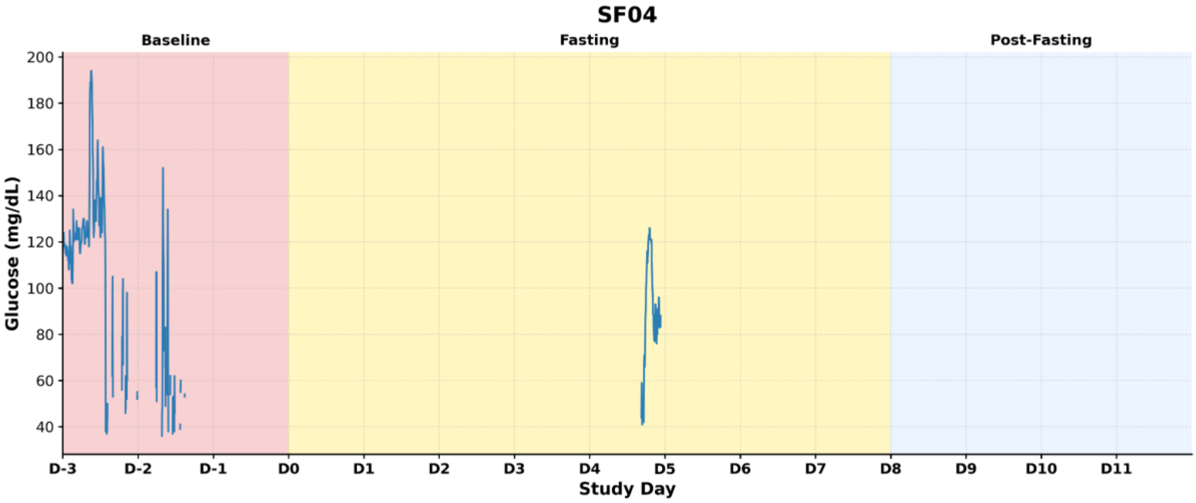


*Figure S1.6 Glucose time-series profile of Subject F04*

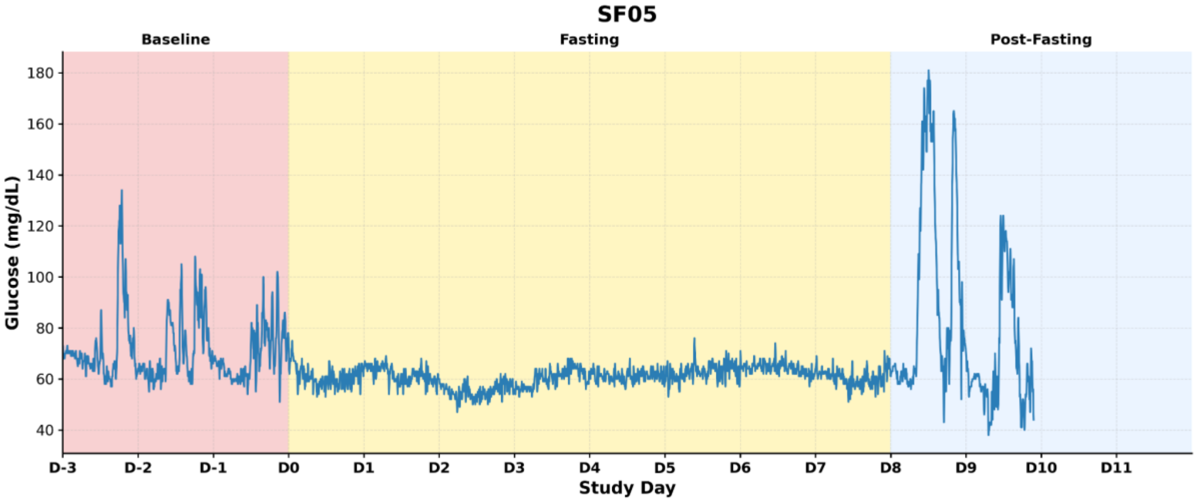


*Figure S1.7 Glucose time-series profile of Subject F05*

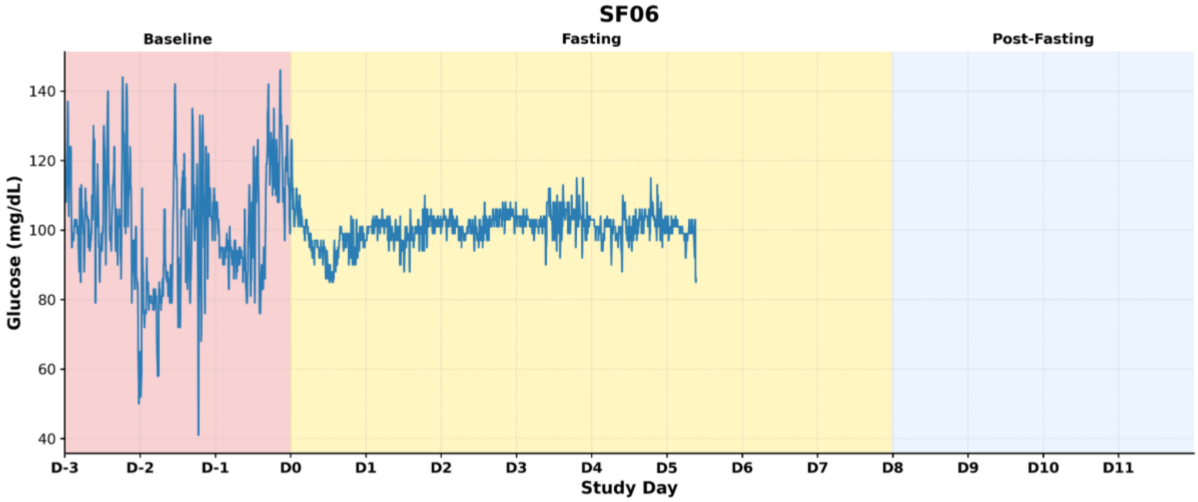


*Figure S1.8 Glucose time-series profile of Subject F06*

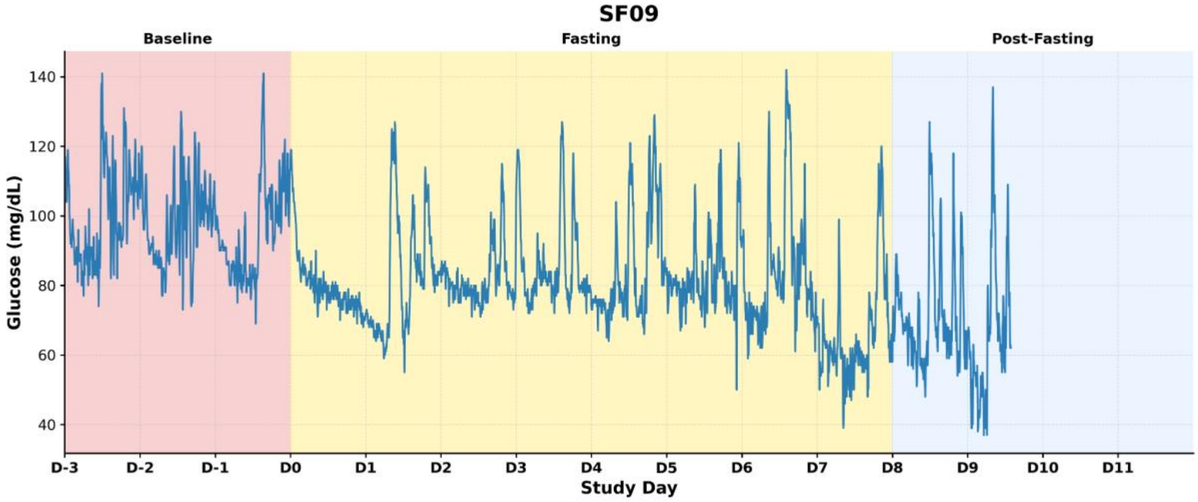


*Figure S1.9 Glucose time-series profile of Subject F09*

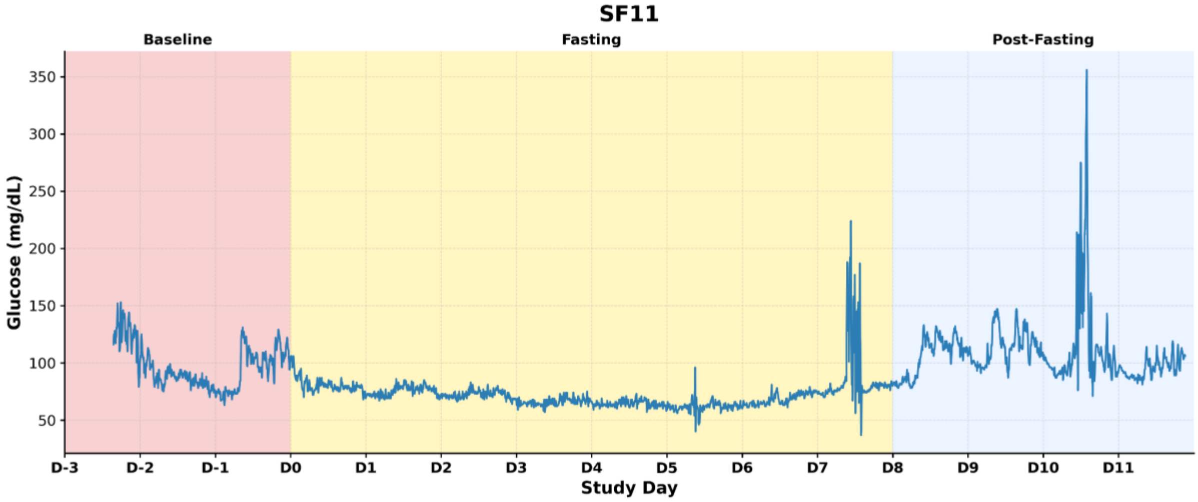


*Figure S1.10 Glucose time-series profile of Subject F11*

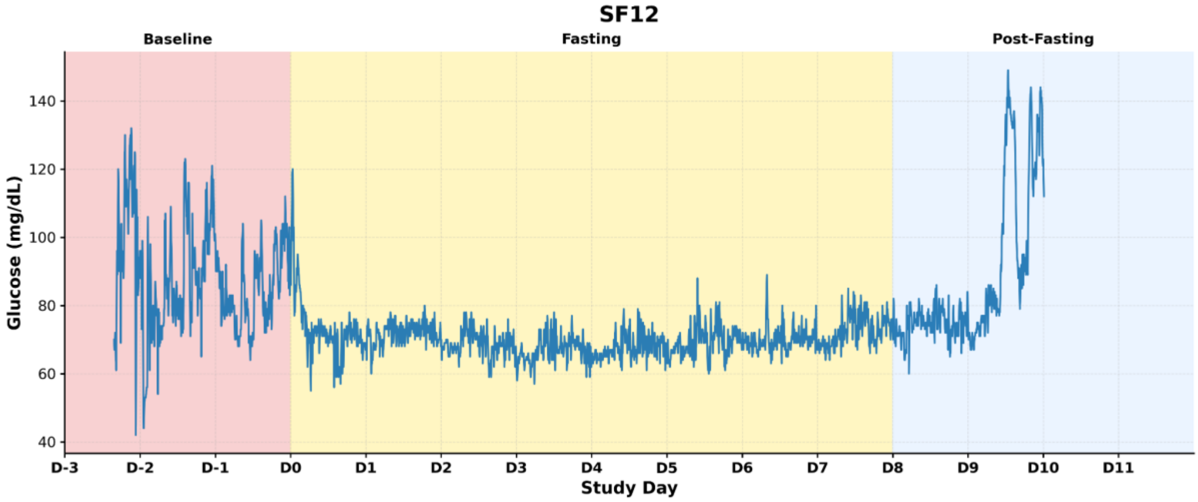


*Figure S1.11 Glucose time-series profile of Subject F12*

**Figure S2. Individual continuous glucose monitoring (CGM) profiles during 30-days fasting.** Raw glucose concentrations (mg/dL) are plotted across study days. Shaded regions represent the baseline (pink), fasting (yellow), and post-fasting/refeeding (blue-grey) phases.

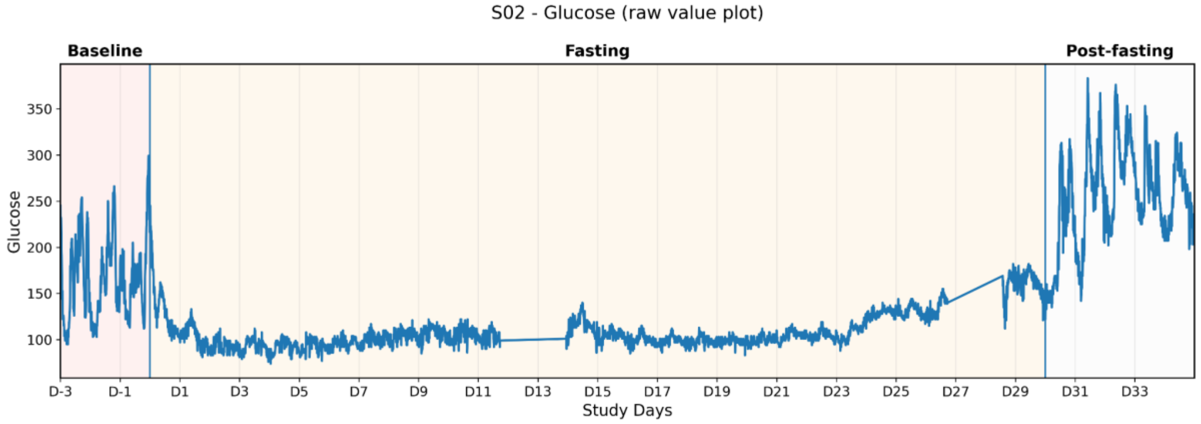


*Figure S2.1 Glucose time-series profile of Subject 02*

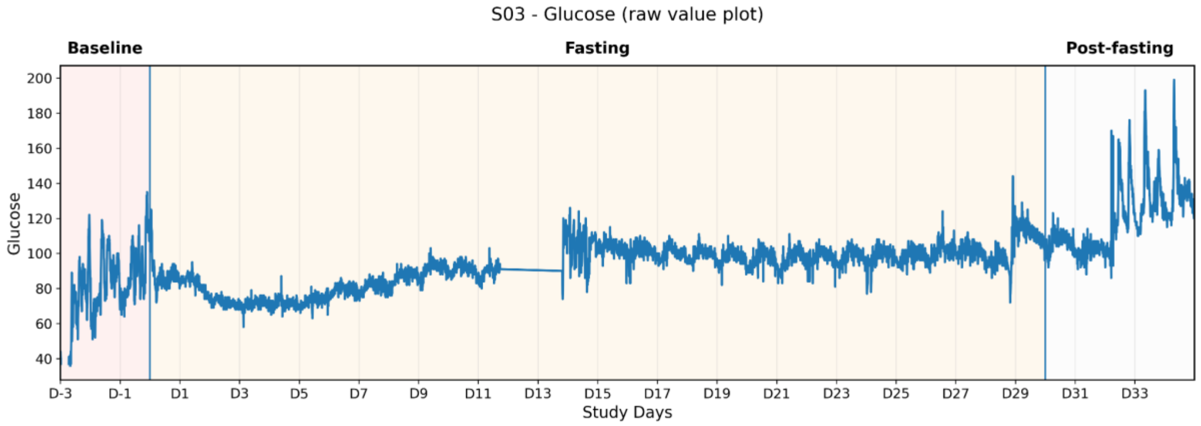


*Figure S2.2 Glucose time-series profile of Subject 03*

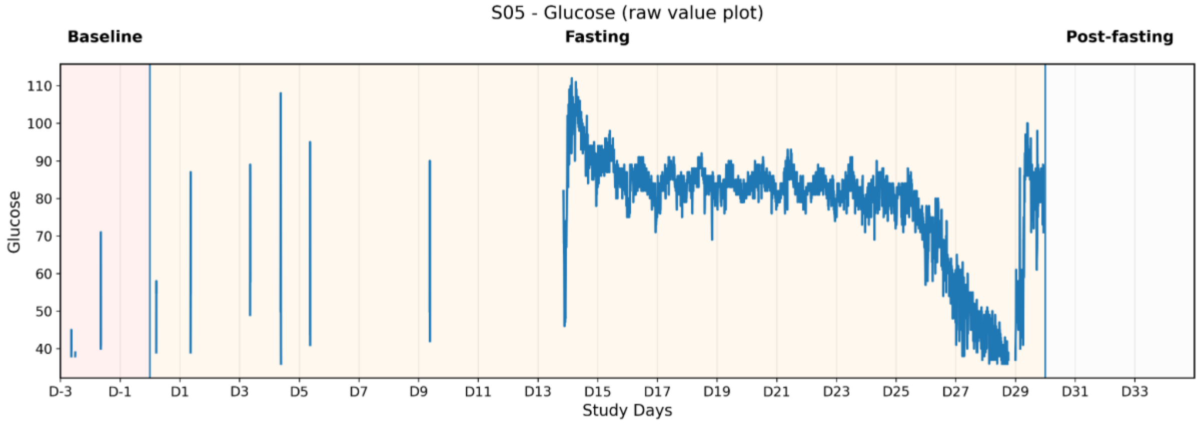


*Figure S2.3 Glucose time-series profile of Subject 05*

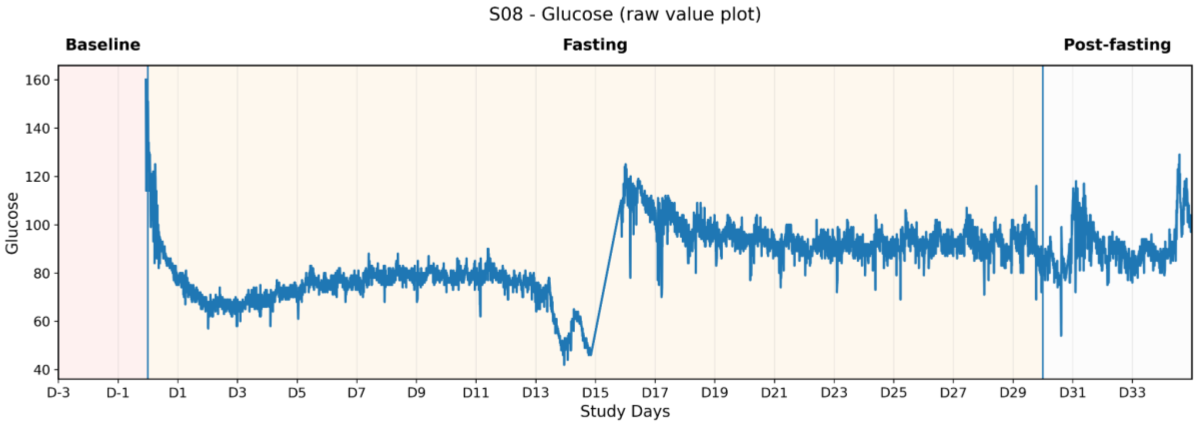


*Figure S2.4 Glucose time-series profile of Subject 08*

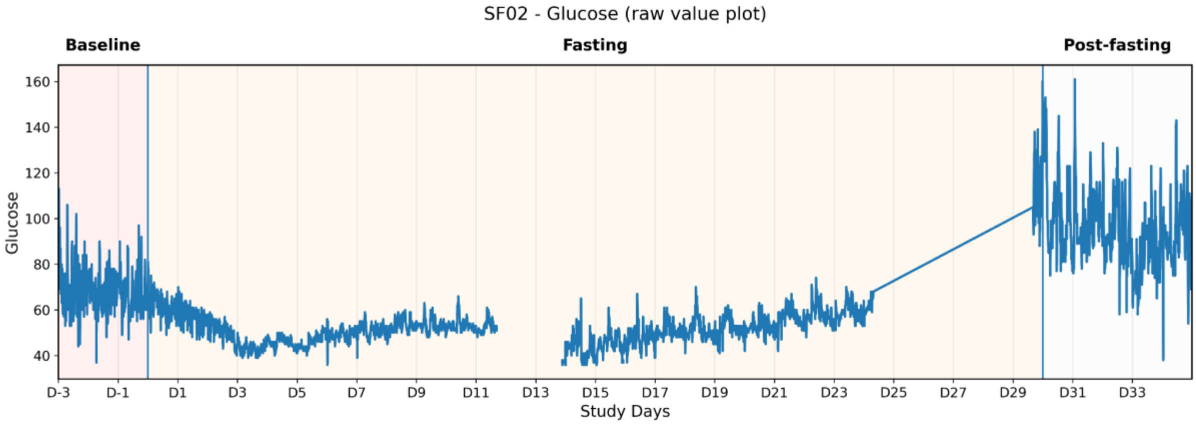


*Figure S2.5 Glucose time-series profile of Subject F02*

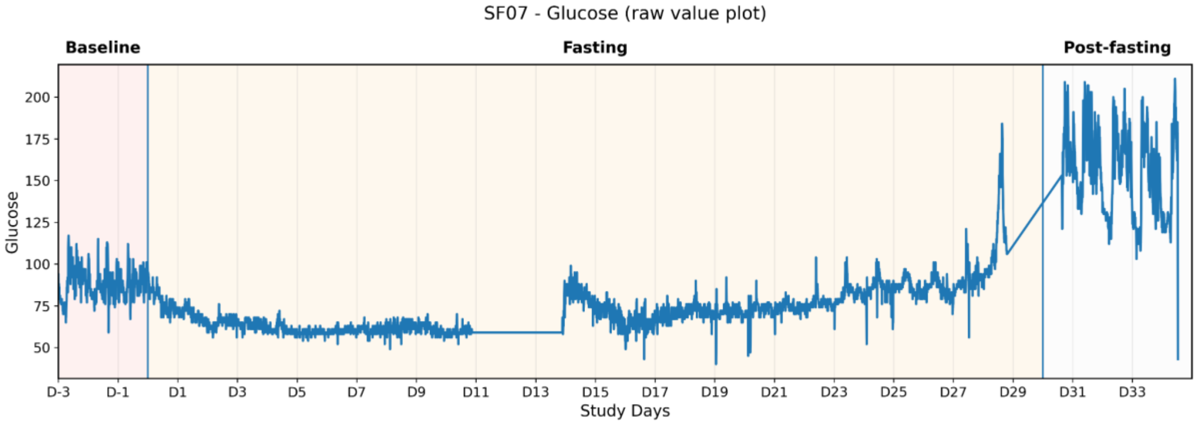


*Figure S2.6 Glucose time-series profile of Subject F07*

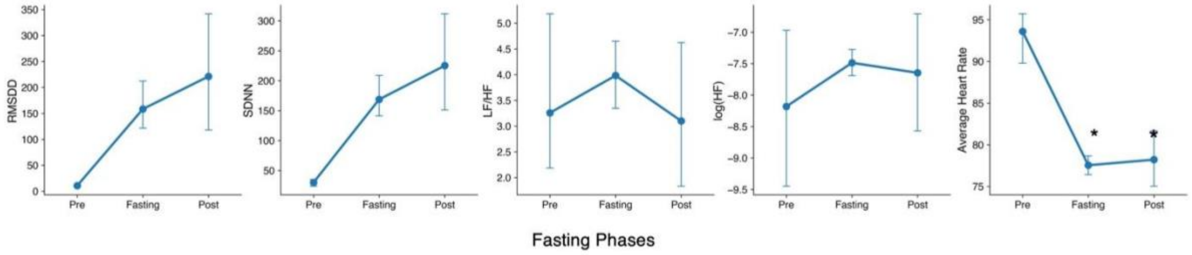


**Figure S3:** HRV parameters (RMSSD, SDNN, LF/HF ratio, logHF and average heart rate) changes during the fasting period. Trends although insignificant show an increase in parasympathetic activity during the fasting period. * denotes statistical significance with respect to baseline with $p < 0.05$.

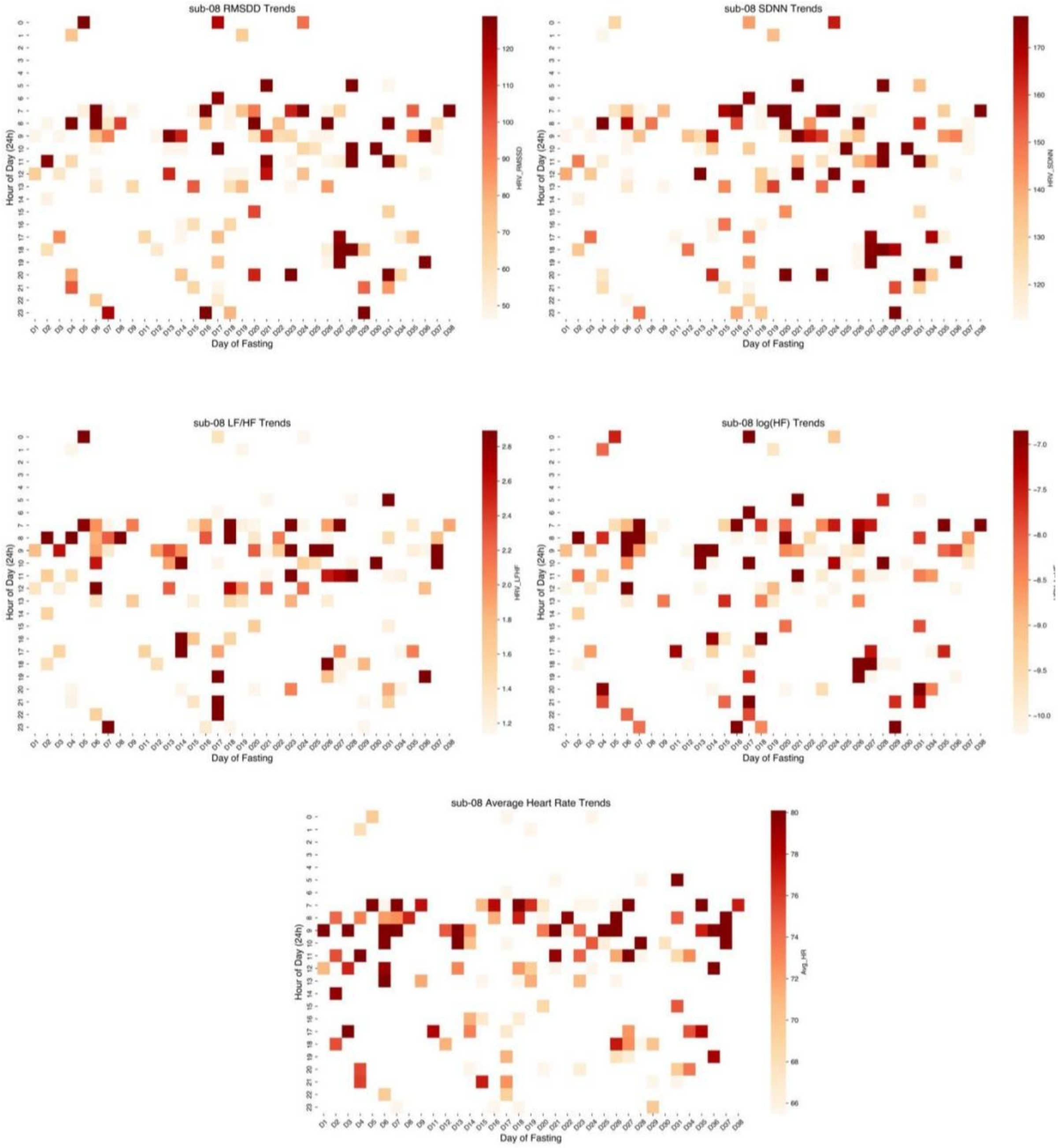


**Figure S4:** HRV parameters variations across different hours of day during the fasting period for sub-08. Here, negative values on the x-axis denotes pre fasting period and greater than thirty would be the post fasting period.
